# Perceptions and Practices of Key Worker Stakeholder Groups in Hospital Animal-Assisted Intervention Programs on Occupational Benefits and Perceived Risks

**DOI:** 10.1101/2020.12.18.20248506

**Authors:** Kathryn R. Dalton, William Altekruse, Peter Campbell, Kathy Ruble, Karen C. Carroll, Roland J. Thorpe, Jacqueline Agnew, Meghan F. Davis

## Abstract

**Background:** Animal-assisted intervention (AAI) programs, used widely for patient benefit, have increasingly been used for healthcare workers (HCW) to reduce occupational stress. However, there are barriers to these programs which limit their utilization, for both patients and HCW, specifically infectious disease concerns. The aim of the research project is to identify barriers and facilitators to AAI program use for healthcare worker benefit, and determine knowledge, beliefs, and practices regarding infectious disease risk and control policies, in order to understand the contextual parameters of program implementation.

**Methods:** We collected perceptions of key stakeholders involved with hospital AAI programs (HCW and AAI workers) through semi-structured in-depth interviews. We used framework analysis to guide thematic coding, completed independently by three researchers.

**Results:** We interviewed 37 participants. We divided our themes into two topic areas: program use for HCW and perceived infectious disease risk. Use for healthcare workers included perspectives on the benefits for HCW and program barriers and facilitators (specifically collaboration and leadership). Perceived risk included opinions on infection concerns with AAI, thoughts on control measures to reduce this risk, and responsibility for safety during these programs.

**Conclusions:** While significant benefits were reported for HCW, they were limited by administrative barriers and hazard concerns. Facilitators to surmount these barriers are best implemented with collaboration across the hospital and appropriate leadership roles to direct safe program implementation. By addressing these barriers through targeted facilitators in the form of evidence-backed guidelines, AAI programs can be used to benefit both patients and HCW.

## Introduction

The numerous benefits of the human-animal bond have extended into the use of animals in healthcare facilities as an adjunctive therapy for patient wellbeing. These animal-assisted intervention (AAI) programs have been shown to reduce stress, pain, and anxiety in patients (Bert et al., 2016; Kamioka et al., 2014; Tsai et al., 2010). One novel program use is for the benefit of healthcare workers (HCWs), given the critical occupational burden they face from high-demand workloads, and secondary traumatic stress from acute negative work experiences. Such stressors can lead to physical, mental, or emotional symptoms such as burnout, depression, and anxiety (Hall et al., 2016; Pradas-Hernandez et al., 2018). Significantly, these symptoms can influence HCW job satisfaction and performance, which have negative downstream effects on patient care (Hall et al., 2016; Monsalve-Reyes et al., 2018). This indicates a crucial need for HCW stress-reduction interventions, and many hospitals are adopting AAI to address this need. To date, no research has evaluated the effectiveness of AAI as a valid therapy to reduce stress in this vulnerable yet essential worker population. If evidence shows that AAI programs can improve occupational health and wellbeing, this will be a previously undescribed benefit of AAI and further promote the human-animal bond in healthcare settings.

Despite the demonstrated benefits to patients, and potential benefits to HCW, there is still hesitancy in the adoption of AAI programs. At the forefront of these challenges is the concern for potential exposure to and spread of infectious disease agents, a challenge that HCW acknowledge (Linder et al., 2017) and which is particularly relevant during the COVID-19 pandemic. Previous research has demonstrated that patients who interact with therapy animals are at higher risk of exposure to hospital-associated pathogens (Dalton et al., 2020; Lefebvre et al., 2009). This indicates the possibility for individuals involved in AAI to become contaminated, including HCW who can transmit microbes to other patients in their care. While guidelines designed to reduce this infectious disease risk have been developed (Murthy et al., 2015), AAI stakeholders need to be aware of the potential risk and be motivated to deploy these control interventions. Thus, hospital infection control strategies for AAI programs need to be effective yet practical to implement, with engagement from HCW and other key stakeholders. There is currently no research on the infection control beliefs and practices for key personnel who work with hospital-based AAI programs. Understanding key stakeholders’ concerns will inform the development of interventions relevant to real-world hospital conditions and will be foundational to future research in this area.

Therefore, this research aimed to collect perceptions of key stakeholders involved with hospital AAI programs on 1) the use of AAI programs as an efficacious occupational stress reduction intervention and 2) concerns and current practices of infection control during AAI programs, including COVID-19. This qualitative study used interviews to formulate more accurate and contextually relevant data, a process shown to be successful in studies on human-animal interaction, AAI programs, and hospital infection control (Degeling & Rock, 2020; Pedersen et al., 2012; Seibert et al., 2014). The ultimate outcome for this qualitative research project is to guide reduction of potential hazards associated with AAI programs, so that these indispensable human-animal bond programs are sustainable as a validated method to holistically improve human and animal wellbeing

## Methods

### Study Population

The Johns Hopkins Bloomberg School of Public Health Institutional Review Board reviewed all study materials and approved this project. To document and describe the perspectives and opinions of hospital-based AAI programs on issues related to risks and benefits, we conducted in-depth interviews with key stakeholders. Key stakeholders included healthcare workers (doctors, nurses, and other patient-care staff) and AAI workers (volunteer handlers and program directors) who work/volunteer in hospitals with existing animal-assisted intervention programs. All stakeholders were over 18 years old and fluent in English. We identified potential study participants from existing contacts and connections through concurrent research studies. Secondarily, we used snowball sampling to identify additional participants (Sadler et al., 2010). Participants were recruited via an email that introduced the research team and study goals.

### Interviews

A semi-structured interview guide was developed by KRD using programmatic framework analysis, a deductive process using predefined central concepts (Gale et al., 2013). The interview guide was edited by co-authors (KR, RT, JA, MFD), and tested with knowledgeable contacts. The interview questions addressed relevant themes connected to the participants’ experiences with hospital animal-assisted intervention, specifically regarding possible concerns and benefits to healthcare workers.

Interviews took place via an online web-conference software between May to July 2020, due to COVID-19 restrictions. Before the start of every interview, participants gave written consent via an electronic signature; occasionally oral consent was obtained from those unable to provide the electronic signature. All interviews were audio-recorded, with participants made aware of the recording before the start. The interviews were conducted by one of three research team members (KRD, WCA, or PC).

### Data Analysis

Audio-recordings from the interviews were transcribed verbatim with the interviewees’ permission. All transcripts were then coded using a combination of previously established deductive codes and inductive codes that arose from the data, per the programmatic framework analysis guidelines (Gale et al., 2013). Each transcript was coded by at least two research members (all coded by KRD, and WCA or PC coding half each). The researchers’ diverse professional backgrounds facilitated openness to different interpretations during both the interview process and data analysis; the first author has a background in veterinary science and public health, the second has a background in mental health and social work, and the third in social disparities and environmental justice. To ensure rigor and inter-coder reliability, the researchers utilized “dialogical intersubjectivity” or open group discussion among both the three interviewers and the wider co-author team for constant comparison of codes to ensure group consensus (Brinkmann & Kvale, 2015; Saldaña, 2015).

Codes were grouped into major themes and sub-themes (Malterud, 2001) to formulate new concepts on the topics. The final step was to explore how these themes were related to each other. Throughout the analysis, the authors reiteratively returned to the interview texts to check that the evolving themes and sub-themes reflected the meanings conveyed by the participants. Representative quotes from the interviews were selected to best illustrate each theme and/or sub-theme.

## Results

### Enrollment and Recruitment

We completed interviews with 37 participants, which are described by occupation in **Table 1**. Interviews lasted from 25 minutes to 1 hour and 5 minutes, with an average time of 42 minutes. Participants were almost equally split between healthcare workers (51%) and individuals directly involved with AAI programs (49%). Three participants labeled as volunteer-handlers and all AAI program directors were hospital employees.

**Table 1:** Participant Recruitment Job Classifications.

| **Study Population** | **Total** |
| --- | --- |
| Healthcare Workers | 19 (51%) |
| Physicians | 4 |
| Nurses | 6 |
| Child Life Specialists | 3 |
| Rehabilitation Therapists (PT/OT) | 2 |
| Clinical Social Workers and Psychologists | 4 |
| AAI Workers | 18 (49%) |
| Volunteer Handlers | 13 |
| AAI Program Directors | 5 |
| Total | 37 |

### Major Themes

Based on our chosen framework analysis methodology, we planned our interviews to focus on the following two main topic areas: 1) the use of AAI programs for HCW, and 2) perceived risks associated with hospital AAI programs. After data collection, we then organized our themes and sub-themes within each of these two topic areas are shown in **Table 2**.

**Table 2:** Themes and Sub-Themes from Qualitative Data.

| Topic Area 1: Program Use for Healthcare Workers |  |  |
| --- | --- | --- |
| Theme 1.1: Benefits | Theme 1.2: Barriers | Theme 1.3: Facilitators |
| Stress reliever and morale booster | Conflict with routine clinical duties | Importance of appropriate staffing for coverage and leadership |
| Improved job function | AAI priority for patients over staff | Importance of collaboration across the hospital and having program advocates |
| Gateway to other therapy | Infection disease risks<br>*(expanded into Topic 2)* |  |

| Topic Area 2: Perceived Infectious Disease Risks |  |  |
| --- | --- | --- |
| Theme 2.1: Concerns | Theme 2.2: Controls | Theme 2.3: Responsibility |
| Infectious and non-infectious concerns | Targets of control strategies | Individuals in charge of safety during these programs |
| Source or cause of infectious concerns | Effectiveness of control strategies and purposed changes | Effectiveness of the training for individuals doing these programs |
| Not concerned about these programs | Adherence to control strategies and barriers to adherence |  |

## Topic Area 1 – Program Use for Healthcare Workers

The first topic area, selected as a focus *a priori*, was on the implementation of AAI programs for HCW usage. All participants, both HCW and AAI workers, felt that these programs, originally designed as complementary interventions to improve patient wellbeing, could be adapted and used for occupational purposes. Some participants even commented that staff needed these programs more than patients. Within this topic area were opinions in three major themes; 1) benefits to HCW from AAI programs, 2) barriers to HCW AAI, and 3) facilitators to overcome the barriers.

### Theme 1.1: Benefits to HCW

All participants reported that AAI programs could benefit HCW, in ways that were similar and unique to patient benefits. Participants felt that benefits from AAI programs to HCW would be heterogeneous depending on personality and coping styles, as well as job function (better for more stressful jobs such as residents and night-shift workers). Reported benefits to HCW were aggregated into three main sub-themes.

#### Sub-Theme 1.1.1. Stress Reliever and Morale Booster

Similar to sessions for patients, participants felt that AAI programs for staff would be a positive distraction or a break from their regular routine. Terms such as “mindfulness” or “reset” were used to describe this positive distraction. The benefits were reported even after brief interactions, with the therapy animal working “instantaneously.” The most commonly reported effect from AAI programs used for staff was the concept of stress reduction. Both HCW and AAI workers reported that occupational stress in this cohort is a significant problem, and these therapy animals could reduce this stress burden.

HCW: “I think that engaging with a dog in a meaningful way de-stresses people.” Beyond reducing stress, participants felt that these programs bolstered morale in HCW receiving this therapy, both in HCW-specific programs or as bystanders to patient-centered programs. This was reported on a personal level and a group level, in that having a therapy animal visit raised the collective mood in the workplace. It was also mentioned that these positive benefits could reinforce the commitment of the hospital to holistic employee wellbeing.

> HCW: “I would think some of the reasons are not just maybe the immediate effect of having that dog, but some of it’s also morale boosting. It’s maybe an indication that the institution you’re working for cares about things like that, and they’re trying to help you have a better work experience.”

#### Sub-Theme 1.1.2. Improved Job Function

Participants reported that HCW benefited from AAI programs through improved job function. This was through its use as an adjunctive therapy modality in patient care, a unique “tool in the toolbox,” resulting in improved clinical outcomes and facilitated communication with patients. In addition, the positive benefits of stress reduction and morale bolstering in HCW also translated into better workplace performance by creating increased employee engagement and resilience.

> AAI Worker: “We know that if our staff are happier and less stressed, that our patients are as well, that carries over to better patient care.”

#### Sub-Theme 1.1.3. Gateway to Other Therapy

The final benefit to HCW that was observed from participant responses was that therapy animals could serve as a mechanism for broaching stress topics. HCW felt more open and freer to discuss mental health and other workplace stress-related factors with a therapy animal present.

> HCW: “There’s definitely something to that human-animal connection. People feel more comfortable disclosing information, I feel like, when the dog is there.”

In addition, HCW were more likely to utilize other stress intervention modalities, such as professional counseling, if combined with AAI programs. The therapy animals served as both an incentive and a non-threatening bridge to what could be considered an “intimidating” or “unneeded” therapy. When combined, it was reported that these therapy programs would appeal to a greater audience, as well as address needs from a broader range of personalities, coping styles, and problems.

> AAI Worker: “We talk about the dog as sort of like a gateway to some other therapeutic interactions where people might be more open to talking to a specialist if they’re, like, petting the dog while they do it.”

### Theme 1.2: Barriers to Programs for HCW

Despite the reported benefits, not all departments and hospitals were able to use AAI programs for their staff. Many of the stated barriers were the same administrative hurdles that were reported as barriers to program use for patients, such as an insufficient number of trained volunteer therapy animals and handlers. Other issues that were common to patient-use included HCW’s fear of and allergies to the therapy animals. However, there were issues that were specific to AAI use for HCW, broken down into three sub-themes.

#### Sub-Theme 1.2.1. Conflicting Timing and Location with Normal Clinical Functions

The most frequently reported barrier to program utilization for HCW was conflicting priorities to their existing job duties. Many HCW participants reported being unable to find time outside of their patient care responsibilities to focus on wellness initiatives, including pet therapy. Handlers also reported this as a barrier when they would attempt to include HCWs in their sessions. Timing issues dealt with both the difficulty of finding a suitable time for HCW-directed sessions, and how long those visits should last in order to be beneficial and worthwhile. The lack of convenience and accessibility of the location for AAI visits was also reported as a potential drawback.

> HCW: “I can’t remember a time when a particularly difficult day has coincided with a dog being available for me to go visit. It’s not like I get to choose the pet therapy over my work.”

#### Sub-Theme 1.2.2. Prioritizing Patient Needs Before Staff

The final primary barrier to HCW AAI sessions was the concept that many HCW felt these sessions should be used for patients. HCW felt that using these programs for themselves, especially knowing the constrained availability of therapy animal dogs and handlers, would remove this limited resource for patients. This concept was also supported by handlers and AAI program directors, who felt the need to prioritize patients because of individual choices or management pressures.

> AAI Worker: “I found that if I’m walking the dog around the unit, a lot of the staff feel like I’m taking the dog away from their patient.”

#### Sub-Theme 1.2.3. Infection Risk as a Barrier to HCW Program Use

Our *a priori* chosen topic area on infectious disease risk in AAI was found *post hoc* to be a sub-theme within the HCW program use topic area; namely that infectious risk was reported as a barrier to the establishment and sustainability of programs for HCW. As such, this sub-theme is addressed in more depth later (*infra* Topic Area 2).

### Theme 1.3: Facilitators to Programs for HCW

In addition to discussing barriers of AAI programs for HCW, opinions on ways to overcome these barriers were also examined. Many of the facilitators described to increase program use for HCW were also relevant to increase program use for patients. Facilitators were grouped into two sub-themes.

#### Sub-Theme 1.3.1. Importance of Appropriate Staffing

A frequently reported facilitator was the value of having adequate staffing to support AAI programs. This was reported to aid in achieving adequate coverage of clinical duties so that HCW could participate in AAI sessions. The importance of having a dedicated staff member in a leadership position, at the institutional and unit level, to take care of appropriate scheduling and administrative tasks was also stressed as a critical factor for HCW AAI program success.

> AAI Worker: “The great thing about a certain dedicated person would be that that was their primary responsibility would be to provide some level of staff support. I think we could reach a lot more staff that way.”

#### Sub-Theme 1.3.2. Importance of Collaboration and Advocates

The last sub-theme identified to aid hospital leadership and staff in implementing HCW-focused AAI sessions was the concept of collaboration across hospital departments and management, including having advocates to promote the value of these programs for staff. These advocates are described as champions in hospital leadership, but also in the greater community who fund the therapy dogs and staff to run these hospital programs. Advocates were reported to be instrumental in securing hospital “buy-in” and increased collaboration.

> HCW: “I think without a champion, it would not get done … their setting up the protocol, not giving up when they hit barriers, making partnerships with places like legal--I think it wouldn’t get done without them, to be honest.”

## Topic Area 2 – Perceived Infectious Disease Risk

The second topic area that was selected as a focus *a priori* for the interviews was the concept of the risk of infectious disease exposure and transmission, including SARS-CoV-2, during AAI programs. This referred to risks to the patients, HCWs, handlers, and therapy dogs. Interviews also concentrated on opinions of infection control policies in the hospital.

The two topic areas were found to overlap in the data, in that infectious disease risks were stated to be a barrier to program implementation, both for patients and HCW (*supra* <u>Theme 1.2</u>). Within this topic area are opinions in three major themes; 1) perceived concerns in AAI programs, 2) control measures, and 3) safety responsibility.

### Theme 2.1: Concerns

Participants discussed general opinions on hazards associated with AAI programs, and described specific incidences that founded these concerns. Concerns for these programs centered mainly on three sub-themes.

#### Sub-Theme 2.1.1. Infectious and Non-Infectious Concerns

For infectious disease risks, the most common concern was the therapy animal serving as an intermediate vector in the spread of pathogenic microbes between patients, HCW, or even the handlers. While the study focused primarily on opinions regarding infectious disease risk, participants occasionally commented on other hazards, such as phobias and allergies to therapy animals, and dog misbehavior (biting, jumping, etc.). Another non-infectious hazard was the therapy animal handler inadvertently causing distress to the patient (through probing questions or privacy issues), but these latter issues were reported as minimal concerns.

> HCW: “I’m concerned about multiple people are touching the same animal. Whatever the person before me passed on, is it staying on the dog. Is it just like another surface that I can just pick it up off of.”

#### Sub-Theme 2.1.2. Source or Cause of Infectious Disease Risk

Participants commented on what they felt was the likely source or reason for these infectious disease risks. Answers were mixed and included the patients and other individuals, the therapy animal, or the hospital environment, with participants frequently mentioning a combination of all three sources.

#### Sub-Theme 2.1.3. Not Concerned About the Programs

While various concerns mentioned above were expressed, a majority of the participants were overall not concerned about these programs and felt the risk for infectious disease was low. Most participants related this to confidence in the control measures in place and adherence to those controls. Many participants also stated that people are unlikely to get infectious diseases from a dog. These opinions were shared equally between healthcare workers and AAI workers, and across the individual roles within each group.

> AAI Worker: “I don’t see that a dog is any more of a vector than any doctor who walked in the room. I mean, we don’t think doctor bringing in disease, but we all bring in bacteria, germs or whatever. I don’t see the dog as more of a vector just because he likes to roll in the mud.”

### Theme 2.2: Controls

Data were collected on knowledge and attitudes about control strategies in place to minimize infectious disease risk in hospital AAI programs. It was mentioned by both HCW and AAI workers that communication and dissemination of these control strategies and any policy updates are critical to program success and can vary across hospitals and departments. These attitudes were aggregated into three sub-themes.

#### Sub-Theme 2.2.1. Goals and Targets of Control Strategies

A majority of participants responded that the goal for these control strategies was ultimately to protect the patient population, but others mentioned protecting the safety of visitors, employees, volunteer handlers, and the therapy animals themselves. Overall, all participants felt control measures and rules were necessary to reduce infectious disease risk and other hazards. When asked what aspect of the therapy visits (the therapy animals, the patients and other individuals, or the hospital environment) the control measures should target to reduce hazards, most participants felt that all three components needed to be addressed in order to comprehensively reduce risk. For those who did select one component, the hospital environment was the component that participants felt should be targeted. Participants, especially AAI workers, felt, in general, that there were more controls directed towards the therapy animals than towards individuals or the hospital environment.

> HCW: “I think you need to control which patients participate, hand hygiene, and I think the handlers are controlling the cleanliness of the dog, and the hospital environment should be clean.”

#### Sub-Theme 2.2.3. Effectiveness of Control Strategies

Participants were asked how successful the current control strategies were towards reducing infection risk of individuals and therapy animals in AAI programs, and queried regarding measures that could be added to current guidelines. Almost all participants felt existing control strategies were effective as long as they were followed and cited a lack of reported negative incidences as evidence of their effectiveness. Participants described additional measures, above the required guidelines, that they implemented to increase safety during the visits, including spacing out the time and location of visits to minimize patient-patient contact, and additional cleaning of the hospital environment, the dog, and dog items (leash, collar, vest, etc.). Reported improvements to current control measures included increased hand-hygiene signs in the hospital’s main lobby and protocols for post-visit dog and handler infection control.

> HCW: “I think that overall the process that we have has really proven to be very effective and very safe since we’ve had little to no incidents with any of the programs.”

#### Sub-Theme 2.2.4. Adherence to Control Strategies

Participants reported that in general adherence to current infection control strategies was very high. Volunteer handlers especially mentioned that their compliance was high because they did not want to jeopardize their access to the hospital. The measure that was stated to have the most variability of adherence was the hand hygiene of patients, visitors, and staff. Other policies, such as no contact precaution patients and pre-visit dog bathing, were said to be adhered to very strictly. It was reported that these control strategies could occasionally be a barrier to participating in AAI sessions by both therapy animal teams and HCW. Handlers felt that bathing the dog before every therapy session was not always feasible or possibly healthy for the dog.

> AAI Worker: “We’re pretty vigilant. It’s really drilled into us. And it’s a privilege to go there. So, you don’t want to do anything to remove that privilege for yourself or for the others.”

### Theme 2.3: Responsibility

A theme that arose from the data was the concept of who is in charge of safety during these visits, and what training goes into preparing these individuals for that responsibility. Perceptions in this theme were broken into two sub-themes.

#### Sub-Theme 2.3.1. Individual in Charge of Program Safety

Opinions were split between participants on who was responsible. Some stated that the volunteer handler was the person primarily in charge of protecting safety, particularly those programs where the volunteer would see patients without the escort of a hospital employee. Other participants said it was a combined responsibility, where the hospital employee was in charge of the patient and the handler was in charge of the dog. It was also mentioned that hospital leadership (both AAI program directors and hospital administration), as well as the organizations certifying the therapy dog teams, had an important role in the safety of these visits, since they were in charge of designing control measures and ensuring adherence.

> HCW: “I feel like the handler and the therapist have equal responsibility in the safety. My goal is to keep my patient safe, and I think the handler should keep the dog safe, so if they’re both safe we’re good.”

#### Sub-Theme 2.3.2. Training of Handlers and Other Responsible Parties

Since handlers were frequently reported to be in charge of these visits, opinions on the level of training they received to reduce infectious disease risks were reviewed, and how effective their training was in making sure they are comfortable to lead these AAI visits. In general, participants felt the training was adequate, with both the volunteers stating they felt prepared and HCW commenting on the knowledge of volunteer handlers.

> AAI Worker: “Handlers should try to educate themselves about so that they can be advocates for themselves and for their *[animal]* partners to have a safer experience.”

## Discussion

This study evaluated perspectives on risk and benefits in hospital AAI programs from key stakeholders, namely healthcare workers and AAI workers. The qualitative study design, which has been previously shown to be effective at identifying benefits for AAI in patient populations (Shen, Abrahamson 2016), allowed us to obtain knowledge, beliefs, and practices from individuals who are intimately involved in these programs. We found major themes within each of the two topic areas; program use for staff including benefits, barriers, and facilitators, and infectious disease risk including concerns, control measures, and responsibility. These themes link together and can provide insight into appropriate program implementation.

### Perceived Risk influences Program Utilization, for Staff and Patients

The benefits of AAI programs for staff—stress reduction, morale booster, improved job function, and gateway to other therapy—were stressed throughout the interviews by both HCW and AAI workers. Stress reduction and morale bolstering from AAI have been previously shown in adult patient populations (Ein et al., 2018; Waite et al., 2018), however even though HCW face different occupational stressors than patients, they expressed similar benefits. HCW stress reduction interventions, such as mental health counseling, yoga/mediation, and group bonding discussions, have previously been associated with improved job function (Brand et al., 2017; Hall et al., 2016; Pradas-Hernandez et al., 2018), and this study supported this concept for AAI. An interesting finding was how AAI sessions could be combined with other proven therapy programs to have a potential synergistic beneficial effect and be more inclusive of people with different personalities and coping styles. For program implementation, it may be advantageous if therapy dog handlers receive training in basic human stress reduction techniques, which could be applied to patients and HCW.

However, the benefits of AAI programs are limited by the reported barriers – administrative barriers (conflict with clinical duties and patient priority) and concerns of infectious disease risk. Participants highlighted the importance of leadership roles to overcome administrative barriers, both within the department to aid staff scheduling, and at the administration/management level to advocate for inclusion of these programs as an important tool for HCW wellbeing. AAI was reported to be a finite resource, for staff support, number and availability of therapy dog teams, and program funding, which limits its use, particularly for HCW, who would rather it be used for patients. However, with successful advocacy and administrative buy-in, these programs could obtain the support they need to grow, to create a “win-win” situation for patient and HCW wellbeing. Leadership and program advocates could push that HCW involvement be part of the mission statement of hospital AAI programs, and harmonize the benefits for patients and HCW.

The other topic area was risks, primarily infectious disease, related to hospital AAI programs, both for patients and staff. A surprising number of participants reported that they had few concerns for these programs, attributed mainly to the efficacy of (with strong adherence to) control measures. This is reflected in published guidelines from major healthcare organizations that promote AAI therapy as a low-risk activity (Murthy et al., 2015). Yet, there is a drawback to this lack of concern — complacency could lead to improperly applied control measures and create tensions with individuals who do have concerns and hesitancy for AAI, particularly those in positions of leadership and management. The best situation is if individuals on the ground are aware of the risk, understand the magnitude of the risk, and know the appropriate methods to reduce that risk. Previous qualitative studies have shown that HCW knowledge and attitudes towards pathogen transmission precautions affect their implementation and adherence (Nichols & Badger, 2008; Saint et al., 2008; Seibert et al., 2014; Yiwen et al., 2010). It is necessary that staff be aware of and understand existing policies, including their rationale, which relates back to the important role of leadership in proper training and communication. Interestingly, in this study, most participants commented on the low risk of the dogs bringing an infectious agent into the hospital, but only a few talked about the role of the dog as a potential intermediary vector, and even fewer discussed the role of the handler. Research has shown that therapy animals can carry hospital-associated pathogens (Dalton et al., 2020), therefore acknowledging this potential risk, and focusing on ways to minimize it, is critical for the program safety.

Program benefits may be strengthened by understanding potential risk, designing and implementing appropriate control measures, ensuring adherence, and continued monitoring by designated leadership. Infectious disease concerns are one of the major barriers to program utilization for both patients and HCW. This barrier is addressed through control strategies, leadership, and collaboration, which will ensure the continued use and potential expansion of these beneficial programs. Like many other human-animal bond programs, a comprehensive and holistic outlook is needed in order to ensure program sustainability.

### Limitations and Future Directions

While this research has many strengths and innovations, there are a few limitations. The first is that the majority of our participants worked or volunteered at pediatric hospitals, rather than adult hospitals. While AAI programs are more frequently used in pediatric populations (Linder et al., 2017), capturing opinions on adult populations may uncover different perspectives and more widespread findings that could be applied to other settings, such as nursing homes and long-term care facilities. Second, it was recognized there were significant differences in protocols across hospitals, and majority of our participants were from three hospitals. Including more hospitals with heterogeneous programs, staff knowledge and buy-in, and infection control policies may lead to different findings, as well as interviews not conducted online during an active pandemic. The findings from this research can be used to design a wide-reaching quantitative survey, which can capture differences across patient populations and hospital protocols.

The results of this study, and future work in this field, can significantly impact the preservation of hospital-based AAI programs. While it has long been known how beneficial these programs are for patients, their use in HCW populations is a novel application. Given how critical the problem of occupational stress and burnout is to this population, particularly during COVID-19, novel strategies are needed. These foundational results suggest their positive usage for HCW, which potentially could be extended to other high-stress occupations, such as first responders. Evidence-based guidelines that address both administrative and hazard concerns will support safe and effective implementation of hospital AAI programs, and reassure hospital administrations and other leadership roles of the value of the human-animal connection in this setting.

## Data Availability

protected qualitative data

## Acknowledgements

The authors would like to thank Drs. Kaitlin Waite and Sharmaine Miller for their assistance. We would also thank the participants for their cooperation.

## Conflict of Interest

The authors have declared no competing interest.

## Funding

This work has been supported by funding from U.S. Centers for Disease Control and Prevention, National Institute for Occupational Safety and Health to the Johns Hopkins Education and Research Center for Occupational Safety and Health Pilot Project Research Training Award. Indirect funding for this research was supported by the National Institutes of Health, Eunice Kennedy Shriver National Institute of Child Health and Human Development [5R01HD097692-02]. Funding for KRD is provided by a grant from the U.S. Centers for Disease Control and Prevention, National Institute for Occupational Safety and Health to the Johns Hopkins Education and Research Center for Occupational Safety and Health [T42 OH0008428], and the AKC Canine Health Foundation Clinician-Scientist Fellowship 02525-E. Investigators additionally were supported by NIH/Office of the Director (K01OD019918 to M.F.D.).

## Notes

### Author Declarations

The Johns Hopkins Bloomberg School of Public Health Institutional Review Board reviewed all study materials and approved this project.

## References

Bert, F., Gualano, M. R., Camussi, E., Pieve, G., Voglino, G., & Siliquini, R. (2016). Animal assisted intervention□: A systematic review of benefits and risks. European Journal of Integrative Medicine, 8(5), 695–706. https://doi.org/10.1016/j.eujim.2016.05.005

Brand, S. L., Thompson Coon, J., Fleming, L. E., Carroll, L., Bethel, A., & Wyatt, K. (2017). Whole-system approaches to improving the health and wellbeing of healthcare workers: A systematic review. PloS One, 12(12), e0188418. https://doi.org/10.1371/journal.pone.0188418

Brinkmann, S., & Kvale, S. (2015). Interviews: Learning the craft of qualitative research interviewing. Sage Publications.

Dalton, K. R., Waite, K. B., Ruble, K., Carroll, K. C., DeLone, A., Frankenfield, P., … Davis, M. F. (2020). Risks Associated with Animal-Assisted Intervention Programs: A Literature Review. Complementary Therapies in Clinical Practice, 39, 101–145. https://doi.org/10.1101/2020.02.19.20025130

Degeling, C., & Rock, M. (2020). Qualitative Research for One Health: From Methodological Principles to Impactful Applications. Frontiers in Veterinary Science, 7(February), 1–13. https://doi.org/10.3389/fvets.2020.00070

Ein, N., Li, L., & Vickers, K. (2018). The effect of pet therapy on the physiological and subjective stress response: A meta-analysis. Stress and Health□: Journal of the International Society for the Investigation of Stress. https://doi.org/10.1002/smi.2812

Gale, N. K., Heath, G., Cameron, E., Rashid, S., & Redwood, S. (2013). Using the framework method for the analysis of qualitative data in multi-disciplinary health research. BMC Medical Research Methodology, 13(117), 260–261.

Hall, L. H., Johnson, J., Watt, I., Tsipa, A., & O’Connor, D. B. (2016). Healthcare Staff Wellbeing, Burnout, and Patient Safety: A Systematic Review. PloS One, 11(7), e0159015. https://doi.org/10.1371/journal.pone.0159015

Kamioka, H., Okada, S., Tsutani, K., Park, H., Okuizumi, H., Handa, S., … Mutoh, Y. (2014). Effectiveness of animal-assisted therapy: A systematic review of randomized controlled trials. Complementary Therapies in Medicine, 22(2), 371–390. https://doi.org/10.1016/j.ctim.2013.12.016

Lefebvre, S. L., Reid-Smith, R. J., Waltner-Toews, D., & Weese, J. S. (2009). Incidence of acquisition of methicillin-resistant Staphylococcus aureus, Clostridium difficile, and other healthcare–associated pathogens by dogs that participate in animal-assisted interventions. JAVMA, 234(11).

Linder, D. E., Siebens, H. C., Mueller, M. K., Gibbs, D. M., & Freeman, L. M. (2017). Animal-assisted interventions: A national survey of health and safety policies in hospitals, eldercare facilities, and therapy animal organizations. Am J Infect Control, 45(8), 883–887. https://doi.org/10.1016/j.ajic.2017.04.287

Malterud, K. (2001). Qualitative research (en medicina): standards, challenges, and guidelines.: EBSCOhost. Qualitative Research Series, 358(panel 2), 483–488.

Monsalve-Reyes, C. S., San Luis-Costas, C., Gomez-Urquiza, J. L., Albendin-Garcia, L., Aguayo, R., & Canadas-De la Fuente, G. A. (2018). Burnout syndrome and its prevalence in primary care nursing: a systematic review and meta-analysis. BMC Family Practice, 19(1), 59. https://doi.org/10.1186/s12875-018-0748-z

Murthy, R., Bearman, G., Brown, S., Bryant, K., Chinn, R., Hewlett, A., … Weber, D. J. (2015). Animals in Healthcare Facilities□: Recommendations to Minimize Potential Risks. Infect Control Hosp Epidemiol, 36(5), 495–516. https://doi.org/10.1017/ice.2015.15

Nichols, A., & Badger, B. (2008). An investigation of the division between espoused and actual practice in infection control and of the knowledge sources that may underpin this division. British Journal of Infection Control, 9(4), 11–15.

Pedersen, I., Ihlebæk, C., & Kirkevold, M. (2012). Important elements in farm animal-assisted interventions for persons with clinical depression: A qualitative interview study. Disability and Rehabilitation, 34(18), 1526–1534. https://doi.org/10.3109/09638288.2011.650309

Pradas-Hernandez, L., Ariza, T., Gomez-Urquiza, J. L., Albendin-Garcia, L., De la Fuente, E. I., & Canadas-De la Fuente, G. A. (2018). Prevalence of burnout in paediatric nurses: A systematic review and meta-analysis. PloS One, 13(4), e0195039. https://doi.org/10.1371/journal.pone.0195039

Sadler, G. R., Lee, H.-C., Lim, R. S.-H., & Fullerton, J. (2010). Recruitment of hard-to-reach population subgroups via adaptations of the snowball sampling strategy. Nursing & Health Sciences, 12(3), 369–374.

Saint, S., Kowalski, C. P., Forman, J., Damschroder, L., Hofer, T. P., Kaufman, S. R., … Krein, S. L. (2008). A multicenter qualitative study on preventing hospital-acquired urinary tract infection in US hospitals. Infection Control & Hospital Epidemiology, 29(4), 333–341.

Saldaña, J. (2015). The coding manual for qualitative researchers. Sage.

Seibert, D. J., Speroni, K. G., Oh, K. M., Devoe, M. C., & Jacobsen, K. H. (2014). Preventing transmission of MRSA: A qualitative study of health care workers’ attitudes and suggestions. American Journal of Infection Control, 42(4), 405–411. https://doi.org/10.1016/j.ajic.2013.10.008

Tsai, C.-C., Friedmann, E., & Thomas, S. A. (2010). The Effect of Animal-Assisted Therapy on Stress Responses in Hospitalized Children. ANTHROZOOS, 23(3), 245–258. https://doi.org/10.2752/175303710X12750451258977

Waite, T. C., Hamilton, L., & Brien, W. O. (2018). A meta-analysis of Animal Assisted Interventions targeting pain, anxiety and distress in medical settings. Complementary Therapies in Clinical Practice, 33(January), 49–55. https://doi.org/10.1016/j.ctcp.2018.07.006

Yiwen, K., Hegney, D., & Drury, V. (2010). A comprehensive systematic review of healthcare workers’ perceptions of risk from exposure to emerging acute respiratory infectious diseases and the perceived effectiveness of strategies used to facilitate healthy coping in acute hospital and community he. JBI Library of Systematic Reviews, 8(23), 917–971.

